# Primary motor cortex has independent representations for ipsilateral and contralateral arm movements but correlated representations for grasping

**DOI:** 10.1101/19008128

**Authors:** John E Downey, Kristin M Quick, Nathaniel Schwed, Jeffrey M Weiss, George F Wittenberg, Michael L Boninger, Jennifer L Collinger

## Abstract

Motor commands for the arms and hands generally originate in contralateral motor cortex anatomically. However, ipsilateral primary motor cortex shows activity related to arm movement despite the lack of direct connections. The extent to which the activity related to ipsilateral movement is independent from that related to contralateral movement is unclear based on conflicting conclusions in prior work. Here we present the results of bilateral arm and hand movement tasks completed by two human subjects with intracortical microelectrode arrays implanted in left primary motor cortex for a clinical brain-computer interface study. Neural activity was recorded while they attempted to perform arm and hand movements in a virtual environment. This enabled us to quantify the strength and independence of motor cortical activity related to continuous movements of each arm. We also investigated the subjects’ ability to control both arms through a brain-computer interface system. Through a number of experiments, we found that ipsilateral arm movement was represented independently of, but more weakly than, contralateral arm movement. However, the representation of grasping was correlated between the two hands. This difference between hand and arm representation was unexpected, and poses new questions about the different ways primary motor cortex controls hands and arms.

## Introduction

The primary motor cortex (M1) is the final common pathway for most voluntary movement generation, with many corticospinal neurons making monosynaptic connections onto arm and hand lower motor neurons. Anatomical studies in rhesus macaques found that for spinal segments C5-T1, 98% of corticospinal neurons from hand and arm areas of M1 originate from the contralateral cortex, and only 2% originate ipsilaterally (Morecraft et al. 2013). Because contralateral pathways dominate the connection between the arm and the brain, M1 contributes to ipsilateral movement indirectly, and to an uncertain degree.

This uncertainty leaves open the possibility that ipsilateral motor cortex constitutes an independent neural pathway that researchers could target for rehabilitative purposes, such as brain computer interfaces for spinal cord injury and stroke. Consistent with this possibility, there has been evidence suggesting separate neural representations for ipsilateral and contralateral movements, including intracortical recordings in monkeys (Donchin et al. 2002; Steinberg et al. 2002; Cisek et al. 2003; Ganguly et al. 2009; Ames and Churchland 2019; Heming et al. 2019), and electrocorticography (ECoG) (Wisneski et al. 2008; Ganguly et al. 2009; Scherer et al. 2009), functional magnetic resonance (fMRI) (Gallivan et al. 2013), and transcranial magnetic stimulation (TMS) (Brus-Ramer et al. 2009) in humans.

However, there is also evidence that ipsilateral and contralateral representations are correlated, (Cisek et al. 2003; Brus-Ramer et al. 2009; Haar et al. 2017; Bundy et al. 2018; Willett et al. 2019), and may show increased correlation as movement complexity increases (Verstynen et al. 2005; Verstynen and Ivry 2011). In particular, two recent studies involving intracortical recordings in humans (Zhang et al. 2017; Willett et al. 2019) found significant correlation in how movement direction was represented for contralateral and ipsilateral movements. The presence of correlations between ipsilateral and contralateral movement directions could cause decoding errors for BCI users.

In this paper, we first aim to quantify the strength and independence of motor cortical activity associated with contralateral and ipsilateral reaching and grasping. Additionally, we investigate continuous bilateral BCI control using neural recordings from a single hemisphere. Previous work investigated BCI control of two translational degrees-of-freedom per arm (Steinberg et al. 2002; Cisek et al. 2003; Rokni et al. 2003; Ifft et al. 2013). Building on this work, we extend the BCI control movements to include a third translational dimension and a grasp dimension for each arm.

Here we report on data collected from two people with tetraplegia who were implanted with intracortical microelectrode arrays in their left M1 cortices while participating in a BCI clinical trial. The participants performed a series of tasks where they controlled either a left or right robot arm for a block of trials or on alternating trials, or they controlled both arms simultaneously. This data was used to determine the neural tuning for contralateral and ipsilateral arm and hand movements under various types of coordination. Based on previous studies, we expected to find a weaker, independent, representation of ipsilateral arm movements as compared to contralateral movements (Steinberg et al. 2002; Cisek et al. 2003; Wisneski et al. 2008). We also expected that units’ preferred directions for ipsilateral arm movements would shift with the addition of simultaneous contralateral movements (Rokni et al. 2003; Perfiliev 2005; Ifft et al. 2013). The data confirmed both of these expectations. We also expected to find little representation for ipsilateral grasping (Tanji et al. 1988). Instead, we found ipsilateral grasp tuning that was strongly correlated to contralateral grasp tuning (Willett et al. 2019). Finally, using two 88-channel electrode arrays in M1 of a single hemisphere, both subjects were able to control planar movements of two arms independently.

## Materials and Methods

### Subjects

Two subjects participated in this study (NCT01364480 and NCT01894802) conducted under Investigational Device Exemptions from the U.S. Food and Drug Administration and approved by the Institutional Review Boards at the University of Pittsburgh and the Space and Naval Warfare Systems Center Pacific. The participants provided informed consent prior to undergoing research procedures. Subject 1 was a 52-year-old (at time of implant) female with tetraplegia resulting from spinocerebellar degeneration without cerebellar involvement (Boninger et al. 2013; Collinger et al. 2013). Two 96-channel intracortical microelectrode arrays (Blackrock Microsystems, Salt Lake City, Utah) were implanted in the hand and arm region of her left primary motor cortex. Subject 2 was a 28-year-old (at time of implant) male with C5 motor/C6 sensory ASIA B spinal cord injury. Two 88-channel intracortical microelectrode arrays (Blackrock Microsystems, Salt Lake City, Utah) were implanted in the hand and arm region of his left primary motor cortex. He also had two 32-channel microelectrode arrays implanted in somatosensory cortex that were not used in these experiments, but are discussed in (Flesher et al. 2016). Both subjects were right-handed prior to paralysis.

### Neural Recording

Neural signals were recorded using the Neuroport Neural Signal Processor (Blackrock Microsystems, Salt Lake City, Utah). To start each test session a threshold was set at -5.25 times the root-mean-square voltage (RMS) (Christie et al. 2015) for Subject 1, or -4.5 times RMS for Subject 2, on each recording channel. Firing rates for each channel were calculated by counting the number of threshold crossings in each 30 ms bin for Subject 1 or 20 ms bin for Subject 2. Firing rates were then smoothed by low-pass filtering the binned firing rates with an exponential filter. For Subject 1 the filter was 450 ms in duration and for Subject 2 it was 440 ms. Each recording channel was treated as a neural “unit”, producing one firing rate regardless of whether a single neuron or multiunit activity was recorded.

### Experiments

As part of the ongoing clinical trials, both subjects were familiar with using the BCI to control a right-handed virtual or robotic arm using intended movement commands recorded from M1. For this study, the participants completed three experiments in which they attempted to control one *or both* arms (Figure 1A) with varying degrees of freedom: (1) single-arm reaching, (2) alternating reach and grasp, and (3) simultaneous reaching. These experiments were performed using two virtual Modular Prosthetic Limbs (vMPLs, Figure 1B, Videos 1-3). Subject 1 completed an additional experiment during which she controlled two robotic Modular Prosthetic Limbs (MPLs, Figure 1C, Video 4). The primary goals of each experiment were to quantify neural tuning to right and left arm kinematics as well as the subjects’ unassisted BCI performance. Together the experiments provide information regarding the strength and independence of bilateral movement-related activity in left M1, as well as the ability to use this information for BCI control.

**Figure 1.**
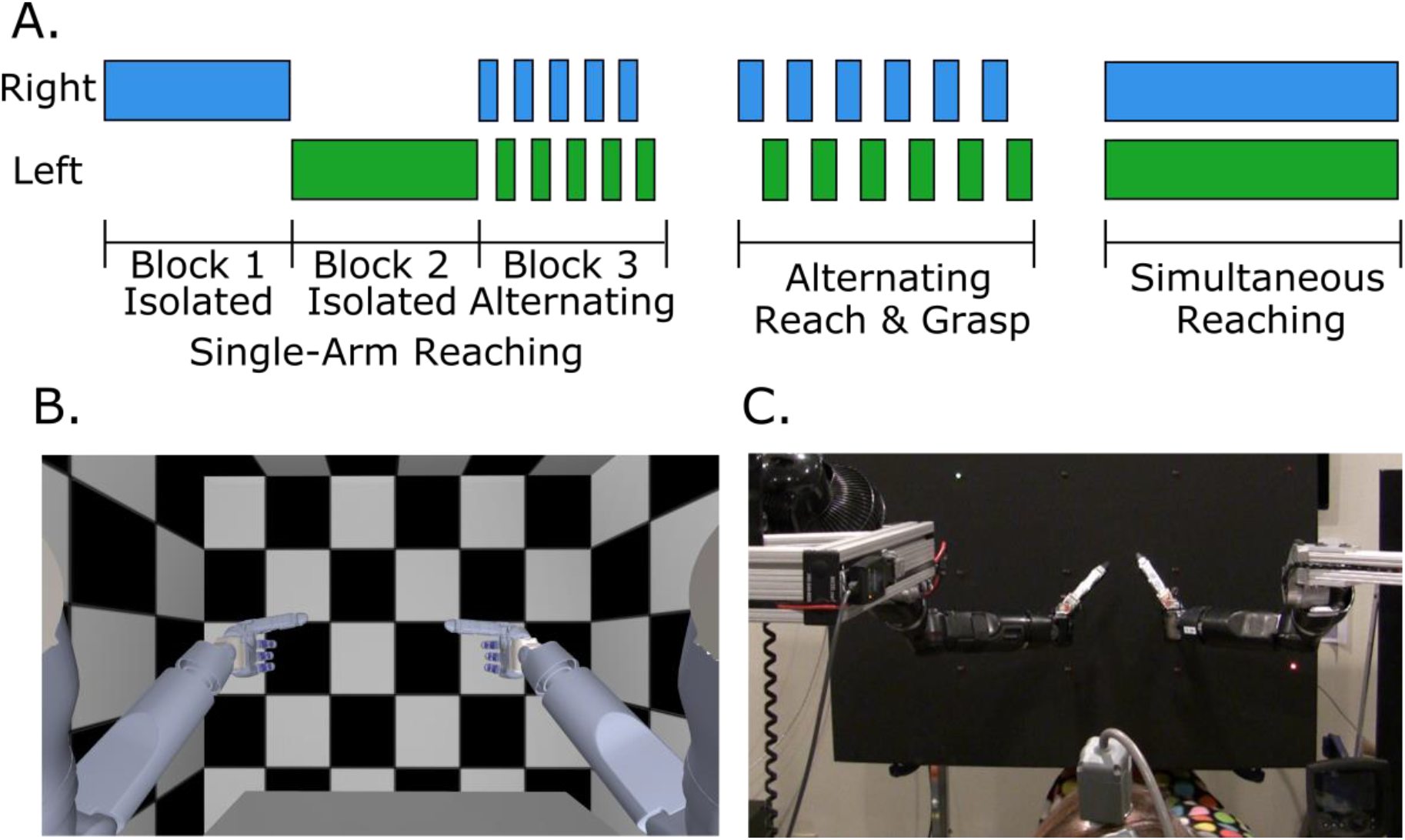
Experiment design and setup. A) Schematics of the three types of experiments showing when right and left arms were used. The first experiment involved solely reaching, where the subject reached with either the right or left arm for the entire block (Blocks 1 and 2). At the end of the experiment, the subject alternated between the right and left arm each trial (Block 3). The second experiment involved reaching and grasping, where the subject alternated between right and left arm after each trial. The third experiment involved simultaneous reaching of right and left arm without grasping. The order of arm use was randomly selected each session. B) The virtual reality environment with the vMPLs. C) A photograph of the physical environment with the two MPLs positioned to either side of the subject. The patient cable and the top of the participant’s head is visible in the bottom center of the image. The left arm’s target is the green light in the upper left, and the right arm’s target is the red light in the bottom right.

### Single-Arm Reaching

#### Overview

The specific goal of this experiment was to evaluate the tuning of motor cortical neurons to 2D reach velocity for the right and left arm. The task consisted of three blocks as shown in Fig 1A. During the first block, the two-dimensional reaching task was completed with either the right or left arm, and then the other arm was used in the second block. The order in which the arms were tested was randomized between days. After both arms were tested in isolation, they were tested together by alternating which arm was controlled for each trial in the third block. Subject 1 completed this task three times between 448 and 453 days post-implantation, and Subject 2 completed this task three times between 367 and 400 days post-implantation.

#### Calibration

A neural decoder was calibrated at the beginning of each block to define the relationship between neural firing rates and movement kinematics to enable BCI control. For each block, the subject viewed two virtual vMPLs (Figure 1B). Reach targets were randomly selected from a set of five locations for each arm. They were arranged in a cross pattern relative to the ‘home’ position shown in Figure 1B. A center target was located at the home position. The left target was 0.18 m from center, the right target was 0.20 m from center, and the top and bottom targets were each 0.23 m from center. The first step for training the neural decoder was to complete a set of observation trials (n=30-60 trials). The subject was instructed to watch the movements and attempt to move the arm towards the targets as they were presented in a random sequence, without returning to the home target between each trial. The vMPL was moved automatically by the computer based on a proportional controller (Video 1). Using neural and endpoint velocity data recorded during observation trials, an encoding model that related the firing rate of a given unit to unilateral arm velocity was computed as:

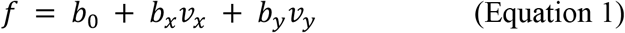

where *f* is the square root transformed unit firing rate, *vx* and *vy* are the velocity vectors of endpoint translational motion in the respective dimensions, and the *b* terms are the coefficients relating the firing rate to the arm endpoint velocity. A unilateral encoding model was fit separately for right and left arm movements using data from Blocks 1 and 2 which contained only unilateral arm movements. Indirect optimal linear estimation (OLE) with ridge regression was then used to create a neural decoder based on the above encoding model (Collinger et al. 2013). For Block 3, when each arm was moved on alternating trials, the encoding model was expanded to include velocity terms for both arms as shown in Eq 2.

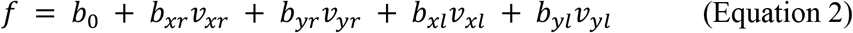

where the subscript *r* indicates endpoint velocities and coefficients that apply to right arm movement, and *l* indicates the terms that apply to left arm movement.

The second step for training the neural decoder consisted of a closed-loop control set with error minimization (n=30-60 trials). During this step, the subject generated velocity commands for the arm using the decoder created from the observation trials, but the computer removed the components of the velocity vector orthogonal to the path to the target (Velliste et al. 2008). The final decoder was then trained using data from the closed-loop calibration set to be used during unassisted brain-control.

#### Brain-control

After each decoder was trained, the subject used it to complete the task without computer assistance before moving on to the next block of the task, which began by calibrating a new decoder (Video 2). Reach targets were presented as described above with a tolerance threshold of 0.05 m around each target. For a trial to be successful, the hand had to reach the target within the defined time limit of 10 seconds or less. Success rates were calculated for each set of trials. During the alternate reaching block (Block 3) the hand that did not have an active target had its tolerance threshold increased to 0.08 m around its previous target, requiring the subject to keep the untargeted hand relatively still.

### Alternating Reach and Grasp

This task was an extension of the single-arm reach experiments that included an additional dimension of arm translation and a dimension of grasp for each arm. Translation targets now included two vertical planes of five virtual target objects each. Once a virtual object target was reached, the hand was required to grasp the object. At that point, a new target appeared and the hand moved the grasped object to the new target and released the object (Video 3). This task was run only under the ‘observation’ condition where subjects observed the computer controlling the arms and attempted to execute the observed movements. The hand-in-use alternated each trial between the left and right hand. We assessed neural tuning by fitting the following encoding model that adds terms for z-translation (i.e., movements towards and away from the subject) and grasp velocities to Equation 2:

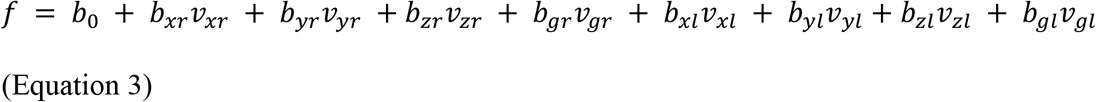

This encoding model was used to identify whether each arm and hand could be controlled independently using the same correlation analysis method described above.

Subject 1 completed the alternating reach and grasp task 6 times between 418 and 439 days post-implantation, and Subject 2 completed the task 4 times between 428 and 448 days post-implantation.

### Simultaneous Reaching

Subject 1 completed a bilateral simultaneous reaching task to test the independence of the control signals for each arm. Two MPLs were used for this task, one on either side of the subject (Figure 1C, Video 4). At the start of each trial, a board in front of the subject simultaneously presented two LED targets. The target for each arm was cued by a different LED color. Both arms moved in the xy-plane (parallel to the board), resulting in four simultaneous dimensions of control. For each hand, the six possible targets were arranged as two columns and three rows. The columns were separated by 22 cm. The top and middle rows were separated by 18 cm, and the middle and bottom rows were separated by 20 cm. The subjects were given a maximum of 10 seconds to successfully complete each trial which involved simultaneously positioning both hands within 8 cm of their respective targets. Decoders were trained using the observation and closed-loop control steps described in the single-arm reaching task, but with the MPLs instead of the vMPLs, and used the encoding model in Equation 2. Using the two MPLs, Subject 1 completed the simultaneous reaching task five times between 490 and 501 days post-implantation. For Subject 2, we only had access to a single MPL and thus were unable to complete this task with him.

### Neural encoding of movement

For each unit, we computed the accuracy (R^2^) of the 2D velocity encoding models for each arm using observation data collected for each block of the experiment. Units that had an R^2^ > 0.005 (all p<0.05) were considered ‘tuned’, and units with an R^2^ > 0.05 were considered ‘well-tuned’.

The preferred direction vector (PD) for each unit during a particular test condition was calculated as the normalized vector of the directional tuning coefficients from Equation 1:

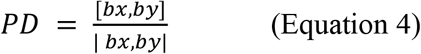

We computed the correlation in preferred direction for the recorded units between right and left arm movements, both when the movements were made in isolation (Blocks 1 and 2) and when they were made on alternating trials (Block 3). The Pearson’s correlation (r) between each dimension of the preferred directions for right and left arm movements was taken across all channels and sessions. We used a randomization test to determine significance of the observed correlations between preferred direction dimensions. After calculating the correlation coefficients for each session, the unit assignments were randomly shuffled within each dimension. We then found the cross correlation for each dimension with the units shuffled to quantify random correlation. This process was repeated 100,000 times for each session and we set R^2^ thresholds at the 95^th^, 99^th^, and 99.9^th^ percentile values.

### Kinematic Performance Analysis

We calculated completion times for BCI-controlled trials as a metric of the quality of the arm control. Trials where the subject was not successful after 10 seconds were not used for these calculations. Additionally, trials were omitted from the single-arm reaching task analysis if they had a very short reach distance. This could occur if the same target as the previous trial was repeated, in which case the arm did not have to move, or if the Euclidean distance to the target was short (< 0.08 m). The latter case could occur due to a buildup of error in the virtual environment, which was rare. Subject 1 had 90 (15%) repeated targets and 18 (3%) short reaches resulting in those trials being omitted from analysis. Subject 2 had 23 (4%) repeated targets and 5 (1%) short reaches omitted. Trials were omitted from the simultaneous reaching task analysis if either of the arms were at the target at the beginning of the trial because of a repeated target (10% of trials). To identify whether the right or left arm could reach the target more quickly, and whether using the arm in isolation was different than using it on alternate trials, trial times were log-transformed to achieve normality and then tested using a 2-way ANOVA for laterality and block of testing.

## Results

### Single-Arm Reaching

#### Bilateral Tuning

Preferred directions were calculated for each unit using the observation data collected at the beginning of each block. The strength of directional tuning was measured as the R^2^ of the encoding model fit (Eq. 2). For both subjects, units were more strongly tuned to right arm (contralateral) movements than left arm (ipsilateral) movements (both subjects, p < 10^−14^, K-S test, Figure 2A). For Subject 1, 70% of units were tuned (R^2^ ≥ 0.005) to right arm movement with 16% being well-tuned (R^2^ ≥ 0.050). For the left arm, 62% were tuned and 5% were well-tuned (out of 576 total recorded units across all days). For Subject 2, 82% of units were tuned to right arm movement with 39% being well-tuned. For the left arm, 75% were tuned and 21% were well-tuned (out of 528 total recorded units).

**Figure 2.**
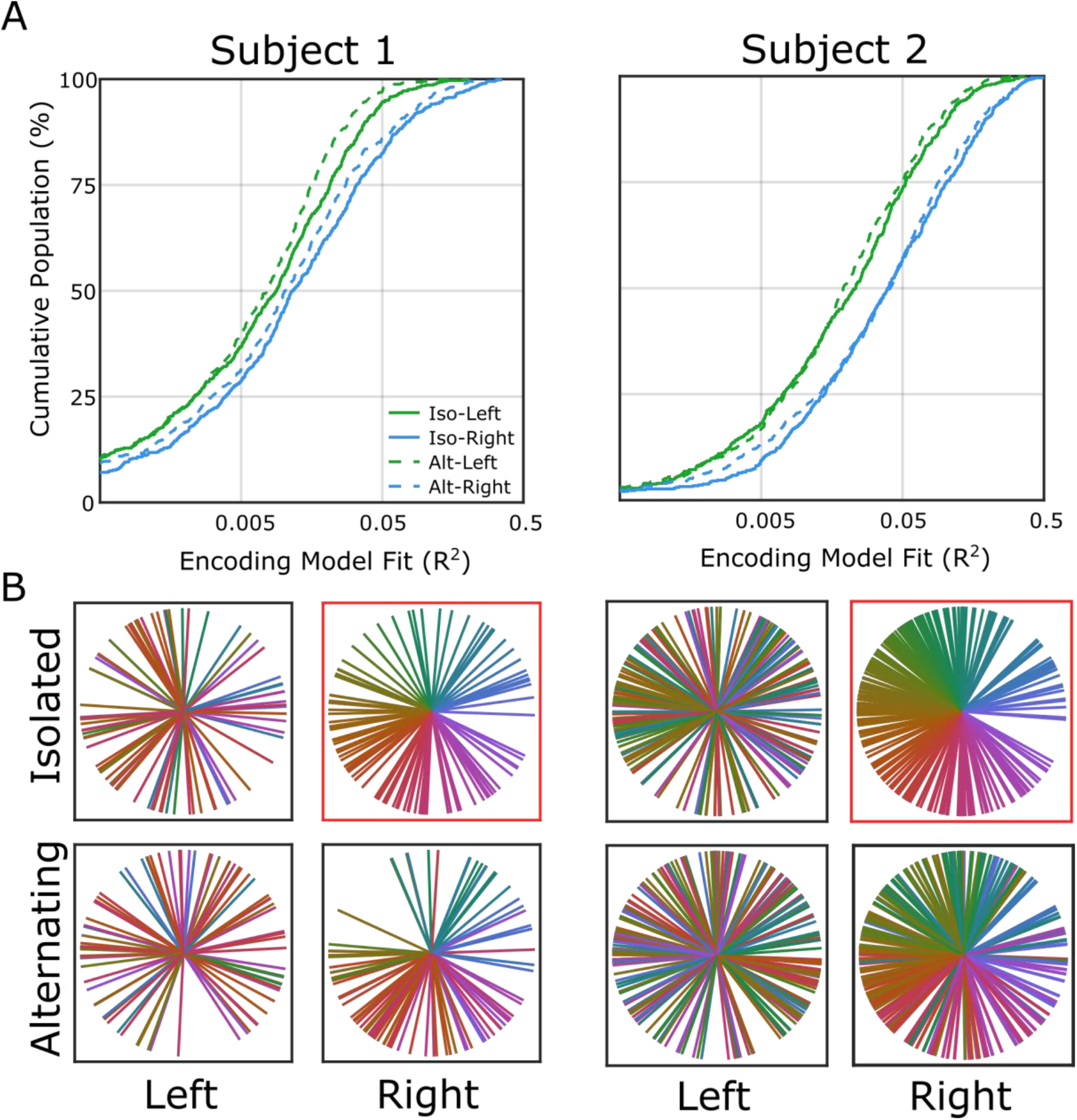
Tuning accuracy and variability across arms and trial types. A) Cumulative distribution plots of the encoding model fit for all recorded units. B) Plots showing preferred directions for units from all sessions that were tuned to both arms during both conditions (Subject 1 on left, Subject 2 on right). Each unit’s color in all four conditions was determined by the angle of its preferred direction during the isolated right arm reaches (in the red box) to allow for visual comparison. Tuning to the right hand is mostly consistent between isolated and alternating reaches, but is not strongly related to left hand tuning (also see Table 1).

In addition to calculating the strength of directional tuning, we also calculated whether the tuning coefficients from the encoding model (Eq. 2) were correlated (Pearson’s r) between arms (left vs. right) and between blocks (isolated vs. alternating reaches). For Subject 1, the tuning coefficients for the right arm were significantly correlated between the isolated and alternating blocks for both x- and y-translation (p<0.001, randomization test, Table 1). Additionally, the tuning coefficients between the right and left arms were significantly correlated during the isolated blocks for x-translation (p<0.01). Notably, that correlation was not present during the alternating block. Subject 2 showed the same three correlations as Subject 1 (all p<0.001). For both subjects, tuning for right hand movement between the isolated and alternating blocks was substantially more correlated than any other comparison, implying that tuning to contralateral arm movement is the most consistent between contexts (Figure 2B, Table 1).

**Table 1.** Preferred direction correlations between arms and trial types for Subject 1 and 2. The Pearson coefficient (|r|) between the two dimensions is shown in each cell. Cells with matching translation dimensions are shaded. Significance shown with * for p<0.05, ** for p < 0.01 and *** for p < 0.001.

| Subject 1 |  |  |  |  |
| --- | --- | --- | --- | --- |
|  | Isolated-Left Arm X | Isolated-Left Arm Y | Alternating-Right Arm X | Alternating-Right Arm Y |
| Isolated-Right Arm X | 0.13** | <0.01 | 0.19*** | 0.02 |
| Isolated-Right Arm Y | 0.06 | 0.01 | 0.04 | 0.31*** |
| Alternating-Left Arm X | <0.01 | 0.01 | 0.06 | 0.03 |
| Alternating-Left Arm Y | 0.03 | <0.01 | 0.01 | 0.07 |

| Subject 2 |  |  |  |  |
| --- | --- | --- | --- | --- |
|  | Isolated-Left Arm X | Isolated-Left Arm Y | Alternating-Right Arm X | Alternating-Right Arm Y |
| Isolated-Right Arm X | 0.17*** | 0.02 | 0.39*** | 0.01 |
| Isolated-Right Arm Y | 0.01 | 0.04 | 0.05 | 0.50*** |
| Alternating-Left Arm X | 0.09 | 0.01 | 0.20*** | 0.02 |
| Alternating-Left Arm Y | 0.04 | 0.14* | 0.07 | 0.03 |

#### BCI Performance

Subject 1 was successful using the BCI to reach with the right arm on 95.8% of trials vs. 52.1% of trials with the left arm. Subject 2 was successful reaching with the right arm on 98.9% vs. 96.7% reaching with the left arm (Table 2). While only Subject 1 showed a detriment in task completion with the ipsilateral left arm, both subjects completed left arm movements significantly slower than right arm movements by 0.70 seconds on average for Subject 1 and 1.44 seconds on average for Subject 2 (both p<10^−8^, 2-way ANOVA). Subject 2 showed no significant effect of block (isolated trials vs. alternating trials) or the interaction between block and arm (left arm vs. right arm trials) on trial time (p = 0.78 and 0.11 respectively, 2-way ANOVA). Subject 1 showed significant effects of both factors (p = 0.03 and 0.01 respectively, 2-way ANOVA) as evidenced by the particularly slow alternating left hand movements (Figure 3).

**Table 2.** BCI control success rates. Success rates are shown for each type of trial, and then combined for all left and all right arm movements.

|  | Isolated-Left Arm | Isolated-Right Arm | Alternating<br>-Left Arm | Alternating<br>-Right Arm | All-Left Arm | All-Right Arm |
| --- | --- | --- | --- | --- | --- | --- |
| Subject 1 | 65.8% | 93.3% | 38.3% | 98.3% | 52.1% | 95.8% |
| Subject 2 | 96.7% | 98.9% | 96.7% | 98.9% | 96.7% | 98.9% |

**Figure 3.**
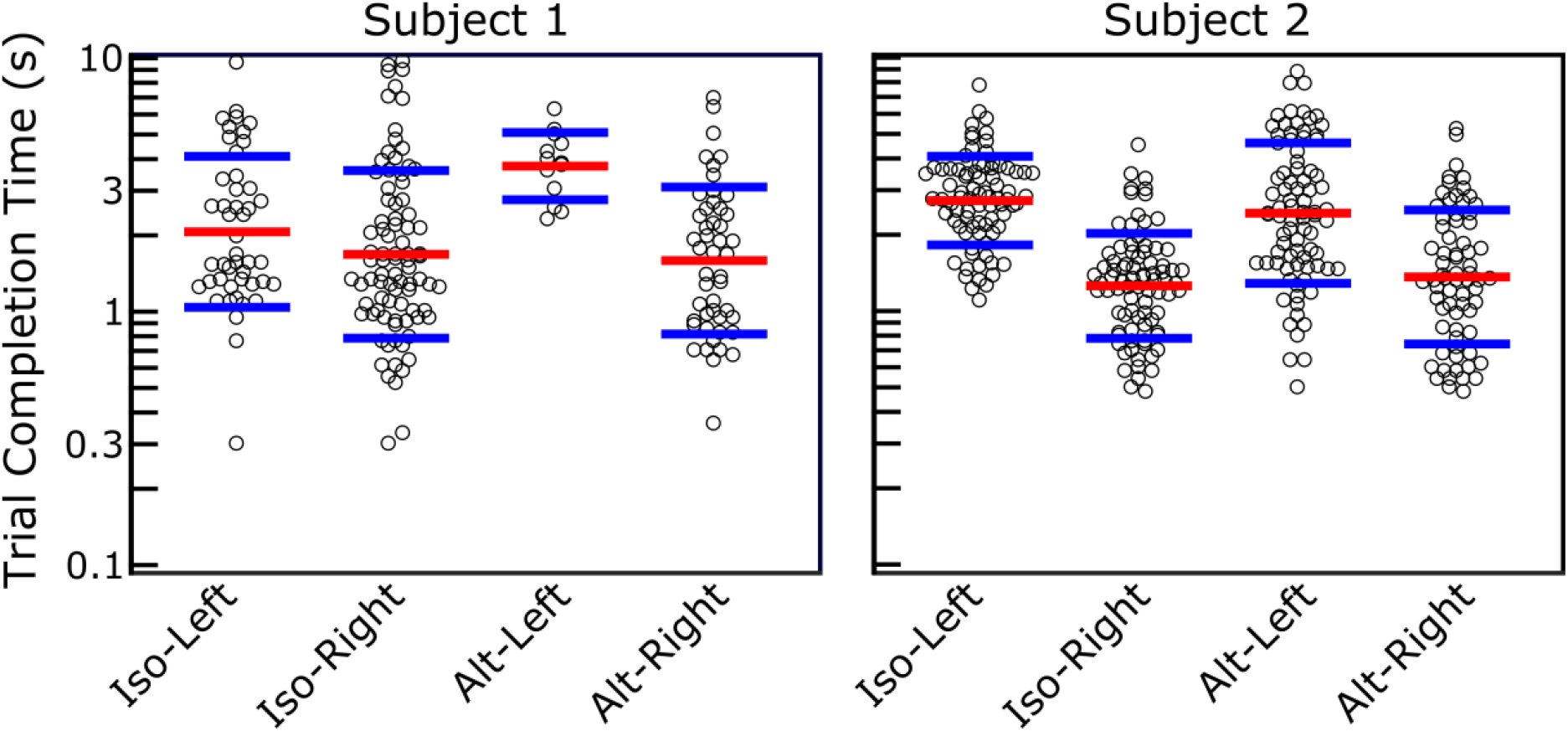
Trial time distributions for single-arm reaching task. Each circle shows the amount of time required to successfully complete a reach, separated by task block and arm used. The red lines show the mean of the log-transformed distribution and the blue lines show one standard deviation. Both subjects were significantly faster in right arm movements (p<10^−8^, 2-way ANOVA).

#### Alternating Reach and Grasp

With this task, we investigated the correlations in neural tuning to grasp and x, y, z translation velocity of the right and left arms. The strongest correlation was between left and right grasping (|r| = 0.47 and 0.36 for Subjects 1 and 2 respectively, Figure 4). Weaker, but significant, correlations for both subjects were noted for different translation dimensions within a single arm. For example, left arm x and y translation were correlated (|r| = 0.24 and 0.22 for Subjects 1 and 2 respectively). The only significant, but still fairly weak, correlation between the two arms during this task was x-translation for Subject 2, matching the results of the reach-only task (|r| = 0.17 and 0.2 respectively, Figure 4 and Table 1).

**Figure 4.**
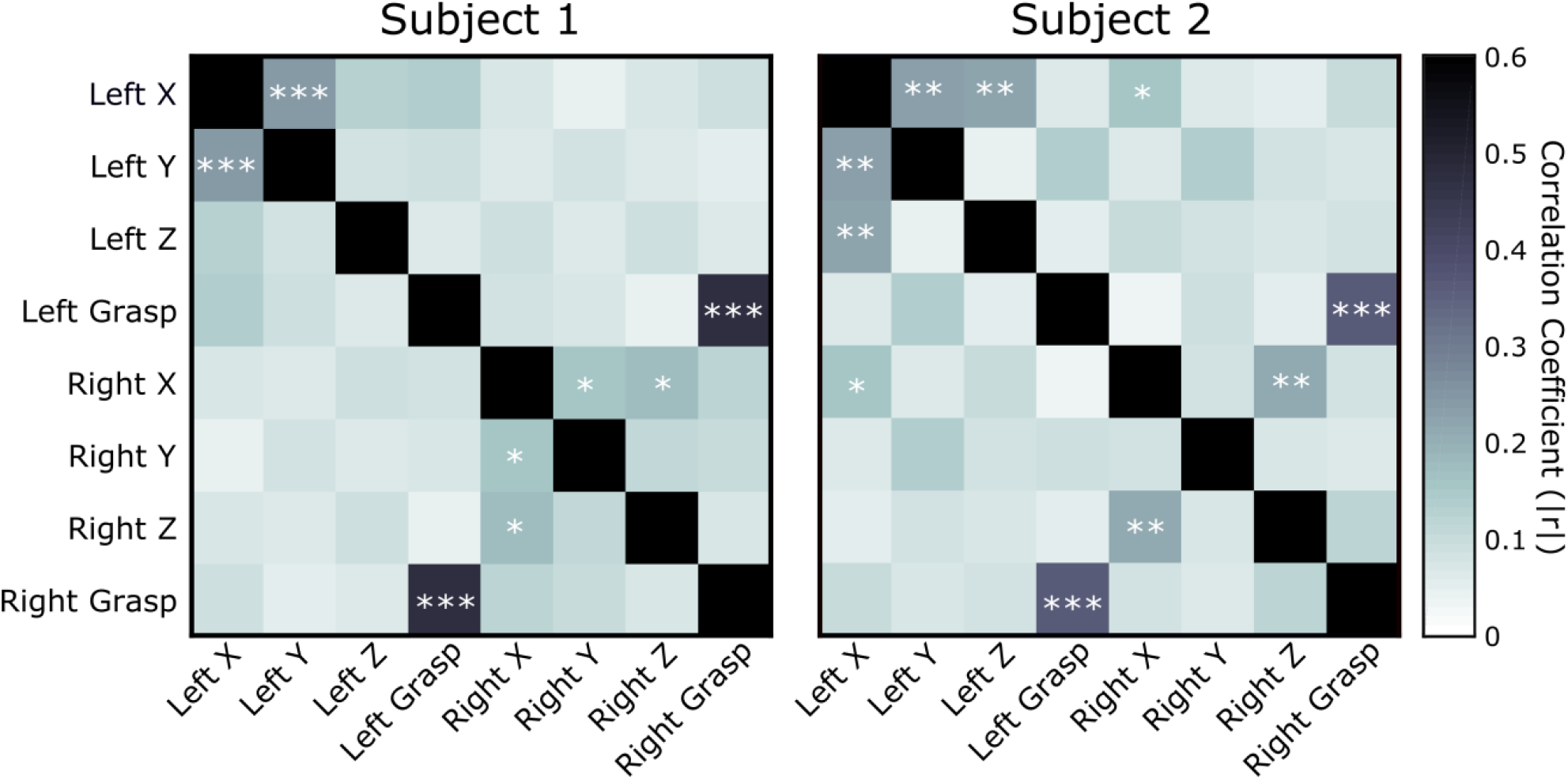
The correlations between preferred directions during the alternating reach and grasp task. The |r| of the correlation is shown, with significance overlaid (uncorrected p < 0.05, 0.01, and 0.001 as *, **, and *** respectively). Right and left hand grasp show the highest level of correlation for both subjects.

#### BCI Control of Simultaneous Reaching

Subject 1 completed simultaneous two-dimensional reaches while seated between the two robotic arms (Video 4). This was the only task completed with physical robotic arms, rather than in the virtual environment. She successfully completed the trials 70.5% of the time (397/563) by positioning both hands in their respective targets at the same time within 10 seconds of target presentation. The right hand failed to reach its target during 2.8% of trials. The left hand did not reach its target during 25.6% of trials. Both hands had to simultaneously be within the target region to be considered successful. Therefore, the subject could fail a trial if the one hand left its target before the second hand entered its target. To further understand this failure mode, we analyzed which hand was outside of its target at the end of the failed trials. At the end of failed trials, the right hand was outside the target 10.8% of the time and the left hand was outside the target 97% of the time. This imbalance in failure modes is consistent with the subject’s verbal report that she needed to attend much more to the left hand’s position, and that when the right hand did not achieve its target it was because she had been distracted by the left hand.

## Discussion

We have shown that the representations for each arm during three-dimensional hand translation are largely independent in unilateral M1. However, grasp tuning for the two hands was highly correlated. This suggests that obtaining independent BCI control signals for grasp would be much more challenging than for arm movements.

Consistent with previous studies in non-human primates (Steinberg et al. 2002; Cisek et al. 2003; Wisneski et al. 2008; Ifft et al. 2013), we found that more channels responded to contralateral, right arm movements than to ipsilateral, left arm movements (Figure 2A). While our analysis was not limited to neurons with direct monosynaptic connections to the arm muscles, our result is consistent with literature suggestion that nearly all of these projections (∼98%) go to the contralateral limb (Morecraft et al. 2013). Therefore, the neural activity associated with ipsilateral movements likely serves a secondary purpose, such as coordination between the limbs (Boisgontier et al. 2014; Willett et al. 2019). One of these secondary purposes has been hypothesized to be inhibition of contralateral arm movement during isolated ipsilateral arm movement (Rokni et al. 2003). However, this is inconsistent with our results that show shifting preferred directions for ipsilateral tuning (Table 1) but no large decrease in the number of tuned units (Figure 2A) that would occur due to general inhibition of the hemisphere. It is also inconsistent with our finding that ipsilateral tuning changes when the arms are used on alternating trials (Figure 2B), and not only during simultaneous use of both arms as reported previously (Rokni et al. 2003; Perfiliev 2005; Ifft et al. 2013). Ipsilateral tuning represents more than a simple inhibition of contralateral movement. The ipsilateral tuning may be related to hemisphere-specific contributions to the type of movement. For instance, studies of patients with unilateral strokes in motor cortex have shown that the left-hemisphere is better at controlling ballistic movements, while the right hemisphere is better at controlling small, accurate movements (Sainburg et al. 2016).

The study participants both had tetraplegia and therefore utilized attempted, but not actual, movements to generate neural activity that enabled control of the BCI. We found that the preferential tuning of M1 activity to contralateral movement led to better BCI control of the right robotic arm than the left arm. Additionally, we found that there was little variability in the preferred directions for each recording channel when computed for the contralateral arm for both unimanual isolated trials and alternating trials (Figure 2B). This is consistent with findings from intact animal experiments showing stable neural activity associated with contralateral arm movements during isolated and bilateral trials (Rokni et al. 2003; Perfiliev 2005; Ifft et al. 2013).

There were several weak, but significant, correlations between control dimensions during the high-dimensional alternating reach and grasp task. These correlations may be the result of not accounting for speed tuning (Moran and Schwartz 1999). Because the correlations were weak, our findings are consistent with studies that find separate neural representations for ipsilateral and contralateral arm movements (Cisek et al. 2003; Ames and Churchland 2019; Heming et al. 2019). However, there was strong correlation between the neural activity generated during left and right hand grasp. This was by far the highest correlation observed for both subjects (Figure 4). This is potentially problematic for BCI and rehabilitation applications, because coordination of bilateral grasping is important for many acts of daily living, such as opening a jar or cutting food, which will be difficult if bilateral grasp information is not independent. There are potential solutions to this problem, such as increasing the number of recorded units to maximize the total available information or making the non-dominant hand grasp controlled in a switch style (Hochberg et al. 2012) to provide stabilization while the dominant hand performs more delicate manipulations.

This study demonstrates simultaneous bilateral BCI control from a unilateral implant in subjects with tetraplegia. Previously, Wisneski et. al. showed alternating one-dimensional BCI control using unilateral ECoG arrays (Wisneski et al. 2008), while Ifft et. al. showed simultaneous four-dimensional control (two-dimensions per arm) as in our task, but with bilateral implants in intact non-human primates (Ifft et al. 2013).

The presence of more well-tuned units for the right arm led to better right arm performance during unassisted trials. Subject 1 showed much better control of the right arm than the left arm while Subject 2, who had more units that were well-tuned to ipsilateral arm movement than Subject 1, showed a smaller, but still significant, advantage in performance for the right arm. Importantly, both subjects showed reasonable left arm control, with Subject 2’s control of the left arm approaching that of the right arm in the low-dimensional task.

One approach to improving this BCI control would be to implant electrodes in both hemispheres of the brain, which has been demonstrated in a primate model (Ifft et al. 2013). However, a single-sided implant would be less invasive. A potential case for a single-sided implant would be a BCI that could enable dexterous control of the contralateral arm while using the ipsilateral arm for holding or stabilizing objects, in the same way that many unilateral amputees use their cable-driven prosthetics (Lunteren et al. 1983; Jang et al. 2011). Additionally, while current intracortical BCI studies are focused on people with tetraplegia (Hochberg et al. 2012; Collinger et al. 2013; Aflalo et al. 2015; Wodlinger et al. 2015; Bouton et al. 2016; Ajiboye et al. 2017), our improved understanding of ipsilateral movement representations could expand motor BCI applications to patients with stroke or upper limb amputation. Stroke patients with severe lesions in one hemisphere’s motor area may be able to receive implants in the opposite intact cortex to use BCI control signals for the otherwise paralyzed arm. This study provides evidence that high-level unilateral arm amputees could benefit from BCI control of a robotic prosthetic limb without disruption from the tuning to the intact ipsilateral arm.

We observed independent tuning of motor cortex activity to three degree-of-freedom end-point translation for both the contralateral and ipsilateral arms, which led to simultaneous BCI control of bilateral reaching. However, the neural activity associated with grasping was highly correlated for both right and left hand movements. This lack of independence could potentially impair the ability of a BCI user to complete bimanual acts of daily living. Future work should investigate the overlap in contralateral and ipsilateral grasp representations to better understand the implications for rehabilitation and BCI development.

## Data Availability

The datasets generated and analysed during the current study are available from the corresponding author on reasonable request.

## Acknowledgements

This study was conducted under Investigational Device Exemptions granted by the US Food and Drug Administration (NCT01364480 and NCT01894802). We thank the participants for their extraordinary commitment and effort in relation to this study and insightful discussions with the study team; Karina Palko for her participation as an honorary research team member and support of the study; Debbie Harrington (Physical Medicine and Rehabilitation) for regulatory management of the study; the University of Pittsburgh Clinical and Translational Science Institute and the Office of Investigator-Sponsored Investigational New Drugs and Investigational Device Exemption support for assistance with protocol development and regulatory reporting and compliance; the volunteer members of the DSMB for their continued monitoring of this study; and Blackrock Microsystems (Salt Lake City, UT, USA) for technical support in relation to this project. This material is based upon work supported by the Defense Advanced Research Projects Agency (DARPA) and Space and Naval Warfare Systems Center Pacific (SSC Pacific) under Contract No. N66001-16-C-4051 and the Revolutionizing Prosthetics program (Contract No. N66001-10-C-4056). The views, opinions, and/or findings contained in this article are those of the authors and should not be interpreted as representing the official views or policies of the Department of Veterans Affairs, Department of Defense, or US Government.

